# Healthcare system overstretch and in-hospital mortality of intubated COVID-19 patients in Greece: an updated analysis, September 2020 to April 2022

**DOI:** 10.1101/2022.09.25.22280326

**Authors:** Theodore Lytras

**Affiliations:** School of Medicine, European University Cyprus, Nicosia, Cyprus

**Keywords:** COVID-19, pandemic, healthcare disparities, intensive care units, right to health, quality of care, intubation, mortality

## Abstract

**Background:** Our previous analysis showed how in-hospital mortality of intubated COVID-19 patients in Greece is adversely affected by patient load and regional disparities. We aimed to update this analysis to include the large “delta” and “omicron” waves that affected Greece during 2021-2022, while also considering the effect of vaccination.

**Methods:** Anonymized surveillance data were analyzed from all COVID-19 patients in Greece intubated between 1 September 2020 and 4 April 2022, and followed up until 17 May 2022. Poisson regression was used to estimate the hazard of dying as a function of fixed and time-varying covariates.

**Results:** Mortality was significantly higher above 400 patients, with an adjusted Hazard Ratio of 1.22, 95% CI: 1.09-1.38), rising progressively up to 1.48 (95% CI: 1.31-1.69) for 800+ patients. Hospitalization away from Attica region was also independently associated with increased mortality, as was hospitalization after 1 September 2021 (HR=1.21, 95% CI: 1.09-1.36). Vaccination did not affect the mortality of these already severely ill patients.

**Conclusion:** Our results confirm that in-hospital mortality of severely ill COVID-19 patients is adversely affected by high patient load and regional disparities, and point to a further significant deterioration after 1 September especially away from Attica and Thessaloniki. This highlights the need for urgent strengthening of healthcare services in Greece, ensuring equitable and high-quality care for all.

## Introduction

During the COVID-19 pandemic, an association between high patient load and in-hospital mortality has been identified in different settings [1, 2]. In Greece, we previously showed how the mortality of intubated COVID-19 patients is affected by regional disparities and patient load, even without exceeding capacity [3]; that analysis did not consider COVID-19 vaccination and only covered the period until May 2021, thereby missing on the large “delta” and “omicron” variant pandemic waves that followed, and which were accompanied by a large number of deaths.

In this context, we aimed to update our analysis in order to (a) validate our previously published findings regarding the association between patient load, regional disparities and mortality of intubated COVID-19 patients, (b) examine whether any changes in this association have happened during the recent period, when “delta” and “omicron” were in circulation, and (c) examine whether vaccination reduces the mortality of already severely ill COVID-19 patients.

## Methods

Our methods have been described before [3]. Briefly, we obtained from the Greek National Public Health Organization (NPHO) anonymized patient data for all cases intubated between 1 September 2020 and 3 April 2022, including vaccination status (number of doses received), dates of intubation, extubation, ICU admission, discharge, outcome at discharge (alive or dead), and followed these cases up to 17 May 2022. No ethical approval was necessary as we used only anonymous surveillance data from which no patient can be identified.

Follow-up time between intubation and extubation or death was split finely into days, and Poisson regression was used to estimate the hazard of dying as a function of fixed and time-varying covariates [4]. Deaths occurring up to five days after extubation were classified as deaths at the end of follow-up. We included in the regression the following covariates: the daily total of intubated COVID-19 patients as an indicator of healthcare system stress, age (as a natural cubic spline with 1 internal knot), sex, vaccination (as a categorical variable), a linear time trend, ICU hospitalization (vs non-ICU), hospital region (the metropolitan regions of Attica and Thessaloniki, vs the rest of Greece), an indicator for the period from 1 September 2021, and an interaction between period and hospital region. Model-based effect estimates were used to calculate Population Attributable Fractions [5] for the different covariates included.

## Results

The series of new intubations, deaths among the study population, and total intubated COVID-19 patients are illustrated in Figure 1. From August 2021 intubations and deaths gradually increased concurrently with the increased circulation of the “delta” variant, and especially so from November 2021. Then after January 2022 circulation of the “omicron” variant prolonged this epidemic wave further, with gradual de-escalation until the end of the study period.

**Figure 1:**
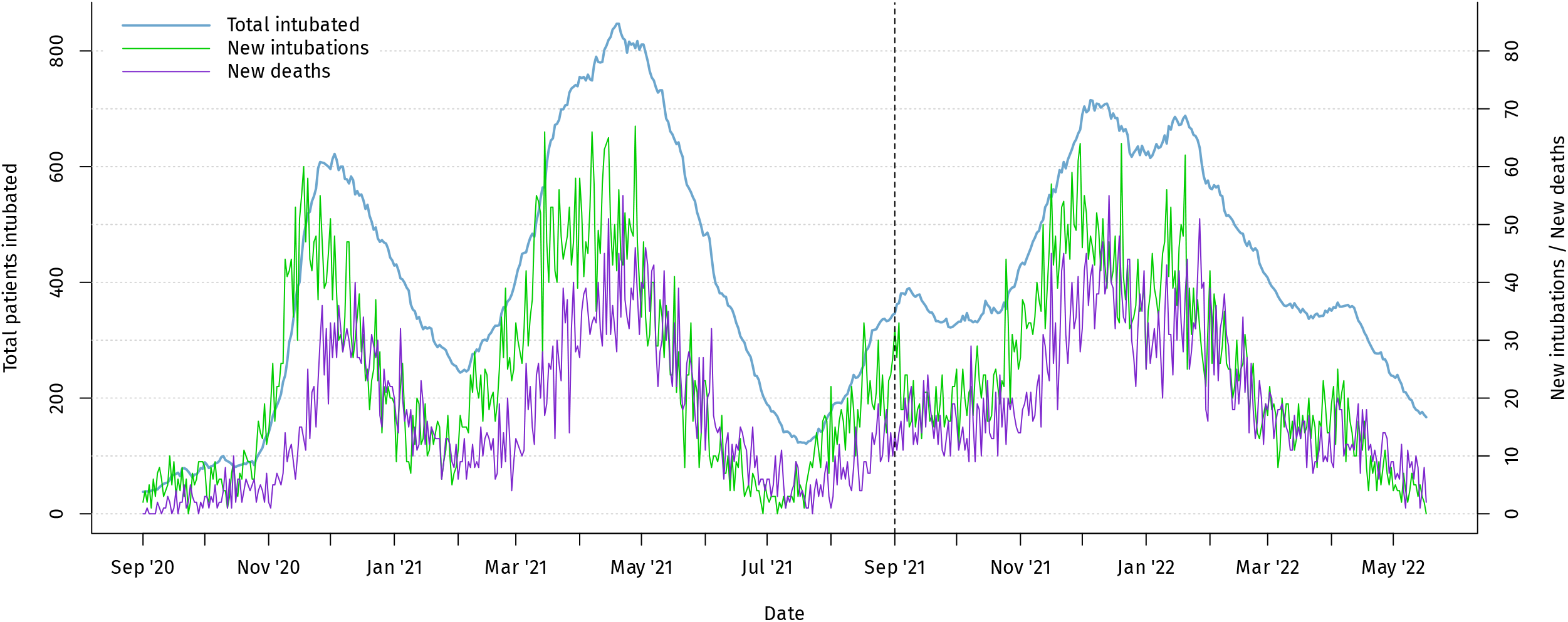
Distribution over time of total intubated COVID-19 patients, new intubations and deaths among the study population, Greece, 1 September 2020 to 17 May 2022.

A total of 14,011 intubated COVID-19 cases were analyzed, of whom 10,466 (74.7%) died (Table 1). Most patients spent part or all of their hospital stay in an ICU (12,902 out of 14,011 patients, with 239,201 out of 250,978 person-days in total). Among those not admitted in an ICU, nearly all died (1084 patients, 97.7%)) compared to 72.7% (9382 patients) among those admitted (p<0.001).

**Table 1:** Characteristics of the study population; laboratory-confirmed COVID-19 cases in Greece intubated between 1 September 2020 and 3 April 2022.

|  | **Total** | **Alive** | **Died** | **p-value\*** |
| --- | --- | --- | --- | --- |
| **Number of patients** | 14011 | 3545 | 10466 |  |
| **Median age, years (IQR)** | 68 (59–75) | 60 (52–68) | 70 (62–77) | <0.001 |
| **Sex** |  |  |  | 0.032 |
| *Female* | 5312 (100) | 1290 (24) | 4022 (76) |  |
| *Male* | 8699 (100) | 2255 (26) | 6444 (74) |  |
| **Vaccination status** |  |  |  | <0.001 |
| *Unvaccinated* | 11944 (100) | 3157 (26) | 8787 (74) |  |
| *Vaccinated with 1 dose* | 574 (100) | 127 (22) | 447 (78) |  |
| *Vaccinated with 2 doses* | 1054 (100) | 175 (17) | 879 (83) |  |
| *Vaccinated with 3 doses* | 439 (100) | 86 (20) | 353 (80) |  |
| **Hospitalization type** |  |  |  | <0.001 |
| *In the ICU* | 12902 (100) | 3520 (27) | 9382 (73) |  |
| *Outside the ICU* | 1109 (100) | 25 (2) | 1084 (98) |  |
| **Time period** |  |  |  | <0.001 |
| *After 1 September 2021* | 6163 (100) | 1280 (21) | 4883 (79) |  |
| *Until 31 August 2021* | 7848 (100) | 2265 (29) | 5583 (71) |  |
| **Hospital region** |  |  |  | <0.001 |
| *Attica* | 5892 (100) | 1817 (31) | 4075 (69) |  |
| *Thessaloniki* | 3065 (100) | 760 (25) | 2305 (75) |  |
| *Rest of Greece* | 5054 (100) | 968 (19) | 4086 (81) |  |
| **Total number of people intubated (at date of intubation)** |  |  |  | <0.001 |
| *0-199* | 767 (100) | 277 (36) | 490 (64) |  |
| *200-299* | 821 (100) | 284 (35) | 537 (65) |  |
| *300-399* | 2824 (100) | 756 (27) | 2068 (73) |  |
| *400-499* | 1648 (100) | 442 (27) | 1206 (73) |  |
| *500-599* | 2055 (100) | 459 (22) | 1596 (78) |  |
| *600-699* | 3300 (100) | 708 (21) | 2592 (79) |  |
| *700-799* | 1686 (100) | 396 (23) | 1290 (77) |  |
| *800+* | 910 (100) | 223 (25) | 687 (75) |  |

\* Mann-Whitney test for continuous variables, Fisher's test for categorical variables

All model results, expressed as adjusted Hazard Ratios (HR) and 95% Confidence Intervals (CI), are shown in Figure 2. There was a significant association between mortality and total intubated patients above 400, with its magnitude increasing progressively: from 1.22 (95% CI: 1.09–1.38) for 400-499 patients, up to 1.48 (95% CI: 1.31–1.69) for 800+ patients. Interestingly, for a given patient load there was substantially increased mortality for the period after 1 September 2021, with a HR of 1.21 (95% CI: 1.09–1.36). Being hospitalized outside the capital region of Attica was also associated with increased in-hospital mortality, especially for the rest of the country (besides Attica and Thessaloniki) and even more so after September 1, with an HR of 1.64 (95% CI: 1.54–1.75). In addition, age was strongly associated with mortality, as was being intubated outside an ICU (HR 2.01, 95% CI: 1.89–2.15). Vaccination did not show a statistically significant association with mortality, regardless of the number of doses received (Figure 2).

**Figure 2:**
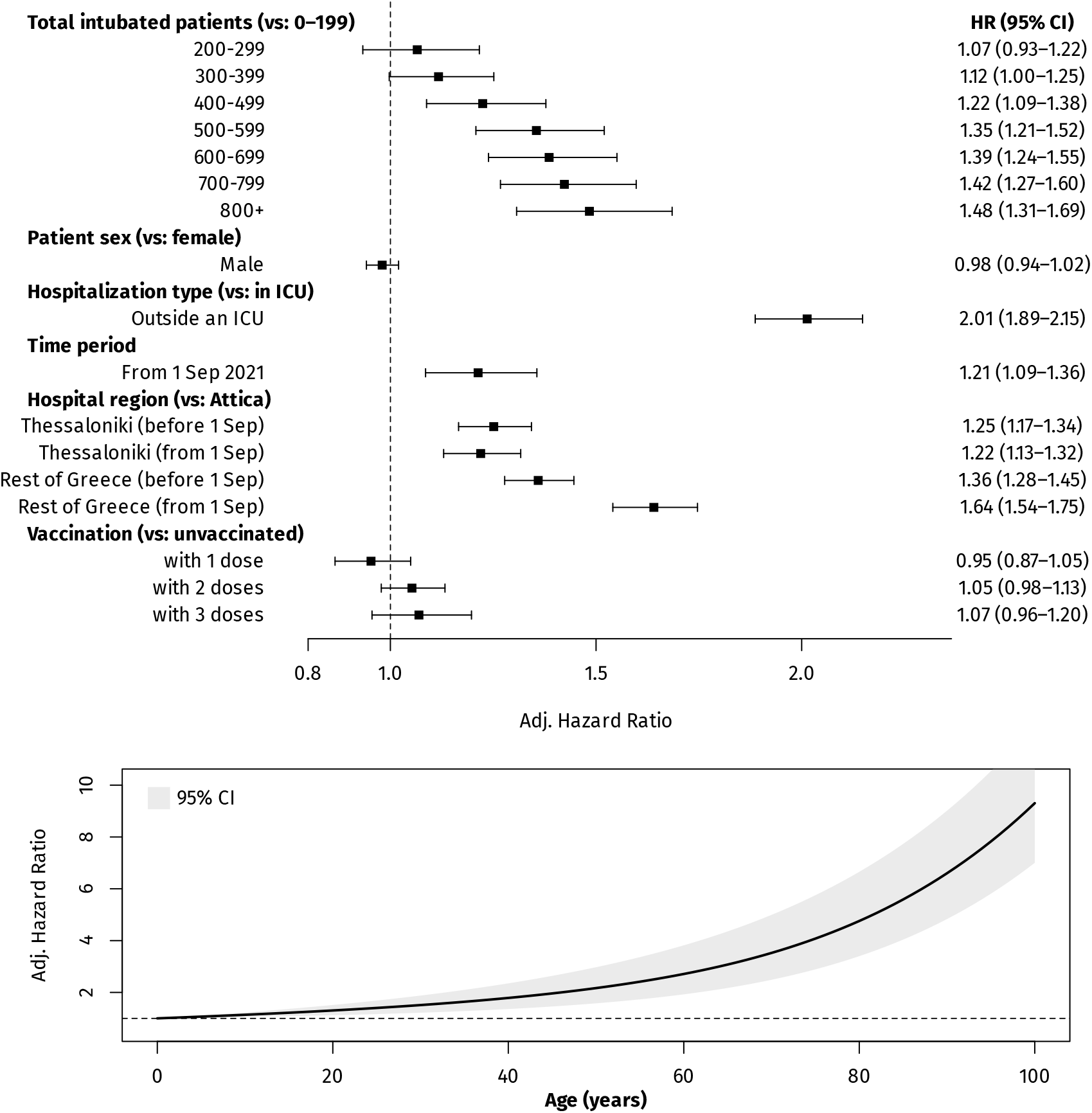
Multivariable (adjusted) associations between in-hospital mortality of intubated COVID-19 patients and age, sex, hospitalization type, hospital region and patient load, Greece, 1 September 2020 to 17 May 2022.

Given the above associations, of the 10,466 deaths reported, 2176 (95% CI: 1297–2986) were attributable to the high load (≥200) of intubated COVID-19 patients, 564 (95% CI: 499–634) to being outside an ICU, 1786 (95% CI: 1572–2002) to being hospitalized away from Attica, and 1196 (95% CI: 779–1586) to being hospitalized after 1 September 2021 and thus experiencing increased mortality. A combined total of 4677 deaths (95% CI: 4021–5284) was attributable to all four factors collectively.

## Discussion

Our analysis confirms the earlier findings of our previously published study, indicating that in-hospital mortality of severely ill COVID-19 patients is adversely affected by high patient load [3]; indeed the associations between patient load and mortality were nearly identical to our previous analysis, but now with significantly higher precision (narrower 95% Confidence Intervals). This shows that patient outcomes are affected not just when healthcare capacity is stressed to depletion, but also at lower levels despite the availability of care not nominally being restricted. This represents a major preventable factor to limit avoidable deaths from COVID-19, and points towards more extensive investment towards preparedness and resilience in healthcare. We also re-confirmed substantial regional disparities, with in-hospital mortality being lower in Thessaloniki and even lower in Attica compared to the rest of the country, highlighting the chronically uneven regional distribution of healthcare resources in Greece [6].

A major new finding is the 21% higher mortality observed in the period starting September 2021, which increases even further for patients hospitalized in the rest of the country (an additional +64%, compared to +36% for the previous period). This suggests that conditions in healthcare services over the past year may have further deteriorated, especially in rural areas (outside Attica and Thessaloniki). It must be noted that from September 2021 the government suspended those healthcare workers that remained unvaccinated against COVID-19 (although many of them eventually got vaccinated and returned to duty, or were replaced with newly hired personnel). Our data cannot prove a causal association between this disciplinary action and the increased mortality, but the temporal coincidence is still worrisome and merits further exploration into the precise causes of this worsening healthcare system performance.

Our analysis reconfirms that being intubated outside an ICU is associated with twice higher mortality than patients inside an ICU, although again this must be interpreted with caution; patient selection (prioritizing for ICU admission those with a higher chance of survival) may account for a large part of this association. On the other hand, we found no statistically association between mortality and the number of COVID-19 vaccine doses received; this suggests that despite vaccination being enormously effective in preventing COVID-19 severe disease and death [7], if a patient has already been intubated as a result of COVID-19 severe disease, it is quality of care and not vaccination that can prevent further deterioration and death.

In fact, the observed non-significant trend towards higher mortality among those having received two or three vaccine doses may be the result of some uncontrolled confounding due to comorbidities, as older people and those with comorbidities tend to be more frequently vaccinated. Indeed, the lack of information on comorbidities and baseline health status is the most significant limitation of our analysis; nevertheless, these factors are unlikely to be differentially distributed over time and with different patient loads, and are both eclipsed by age, which is the major determinant of the risk of death and which we carefully adjusted for [8, 9]. Therefore residual confounding by baseline health is unlikely to have affected these results to any substantial extent.

As a result, the observed associations are most likely to reflect real and avoidable differences in the quality of care for COVID-19 patients due to increased patient load, regional disparities and ICU availability, as well as a true deterioration after 1 September 2021. The findings highlight the need for urgent strengthening of healthcare services in Greece, in order to improve their performance and ensure equitable access to high-quality care for all [10].

## Data Availability

Study data and analysis code are available upon reasonable request to the authors

## Acknowledgements

We acknowledge the staff of the National Public Health Organization for their hard work in coordinating the surveillance of COVID-19 in Greece.

